# Huntingtin Interacting Protein 1 (HIP1) autoantibodies as a novel potential surrogate marker for Rheumatoid Arthritis: Pilot Study

**DOI:** 10.1101/2022.09.07.22279672

**Authors:** Surbhi, Ayushi Goel, Ved Chaturvedi, Sneha Verma, Sonia Rawat, Nirmal Kumar Ganguly, Shivani Arora Mittal

## Abstract

Rheumatoid Arthritis (RA), an autoimmune disease, primarily affects synovial joints but has systemic manifestations upon progression. Considering limited specific diagnostic and prognostic biomarkers, identifying the disease early and monitoring its progression is important. Previous reports have shown that Huntingtin Interacting Protein 1 (HIP1) is over-expressed in rat synoviocytes, and its autoantibodies in sera of some cancers has diagnostic relevance. Here, we explored HIP1 and its autoantibody levels along with Th1/Th2/Th17 cytokines in sera of RA patients for their potential as surrogate markers. Relative level of autoantibodies to HIP1 was detected using an in-house developed ELISA. HIP1 expression was found comparable in RA patients and controls. HIP1 autoantibodies were found significantly raised in RA patients (p=0.002) and were higher in patients with active disease, thereby correlating with disease progression (p=0.042). Elevated Th1 and IL-6 cytokines (p=0.024) were found in a subset of patients with active disease, coinciding with their pro-inflammatory profile. This is the first report demonstrating a humoral immune response against HIP1 in RA patients, correlating with an active disease status. Further studies in a larger cohort are required to validate this as a surrogate marker.

**Key Points:**

⍰ HIP1 autoantibodies are significantly increased in sera of RA patients.
⍰ HIP1 autoantibodies correlate with active disease in RA patients.

## Introduction

Rheumatoid Arthritis (RA) is an autoimmune disease with inflammation and synovitis of peripheral joints as hallmarks. With a global prevalence of 0.5-2%, it has impacted more than 5 million lives, and its incidence is expected to increase in the coming decades [1-2]. Genetics, female sex and environmental factors contribute to susceptibility. Strong links with vitamin D deficiency, smoking, obesity and microbiota have also emerged [3-4]. Clinical management of RA is further complicated by a limited response in some patients to conventional disease-modifying antirheumatic drugs (DMARDs). Recent studies have emphasized the potential of combination therapeutics using conventional and targeted DMARDs [5].

Clinicians have long felt the need to employ a set of biomarkers to improve diagnostic precision in RA. The current diagnostic criteria rely on a combination of serological and radiographic assessments. The most commonly used biomarkers are acute phase proteins such as erythrocyte sedimentation rate (ESR), c-reactive protein (CRP) and rheumatoid factor (RF) [6]. However, they are also non-specific indicators of inflammation [7-8], suggesting the need to identify specific surrogate markers that can aid in early disease identification and monitoring progression. Along these lines, ACPA is considered by far the most precise diagnostic biomarker [9]. However, about 20-30% of RA cases are seronegative and considerably erosive [10].

The genesis of this study stems from previous reports of huntingtin-interacting protein-1 (HIP1) overexpression in mouse fibroblast-like synoviocytes (FLS) [11], demonstration of HIP1 autoantibodies in some cancers [12-13], and commonalities between RA FLS and cancer cells [14-15]. Few previous reports in mice suggested that HIP1 is an essential contributor to RA pathogenesis. Comprehensive profiling of epigenetic signatures in mouse FLS cells identified ‘Huntingtin Disease signaling’ as one of the key enriched RA-specific pathways [16]. Further, HIP1, one of the key proteins of this pathway, is over-expressed and found to increase the invasive abilities of mouse RA FLS cells [12]. HIP1 overexpression is also reported in gliomas, prostate and other epithelial cancers [12,17-18], and its role in increasing cancer cell survival and receptor tyrosine kinase (RTK) signaling is also well established [19]. The presence of autoantibodies to HIP1 in sera is reported in some cancers like gliomas, lymphomas and prostate [13, 17, 20]. In prostate cancer, these are found to be highly specific and are demonstrated as an important marker in combination with others [13]. Moreover, RA FLS and cancer cells have several common features like inflammatory phenotype, transmigration ability and angiogenesis [21-22]. Thus, the aim of our work was to assess the prevalence of HIP1 and its autoantibodies in sera of RA patients and to evaluate its potential as a biomarker. To our knowledge, this is the first study exploring the role of HIP1 in RA patients.

## Results

### Demographic Details of Patients and Controls

Table 1 shows the average age distribution, gender and disease duration of RA patients (n=72) and healthy control group (n=54). Details of the subgroups, consisting of 27 mild and 45 moderate-to-severe RA patients, are shown in Table 2.

**Table 1.** Demographic features of Healthy subjects and RA patients.

| Table 1. Demographic features of Healthy subjects and RA patients |  |  |
| --- | --- | --- |
| Characteristics | Healthy Subjects (n=54) | RA patients (n=72) |
| Female, n (%) | 18(33) | 67 (93) |
| Age (years) Mean ± SD [min, max] | 49.31 ± 11.52 [76, 31] | 49.30 ± 10.57 [70, 31] |
| Disease duration (years) (Mean ± SD) [min, max] | - | 6.33 ± 5.84 [0.08, 30]<sup>1</sup> |
| Rheumatoid Factor (RF), n (%) positive) | - | 20 (69)<sup>2</sup> |
| Anti-cyclic citrullinated peptide (anti-CCP), n (%) |  | 17 (73)<sup>3</sup> |
| <sup>1</sup>Data were available for 61 patients only<br><sup>2</sup>Data were available for 29 patients only<br><sup>3</sup>Data were available for 23 patients only |  |  |

**Table 2.**
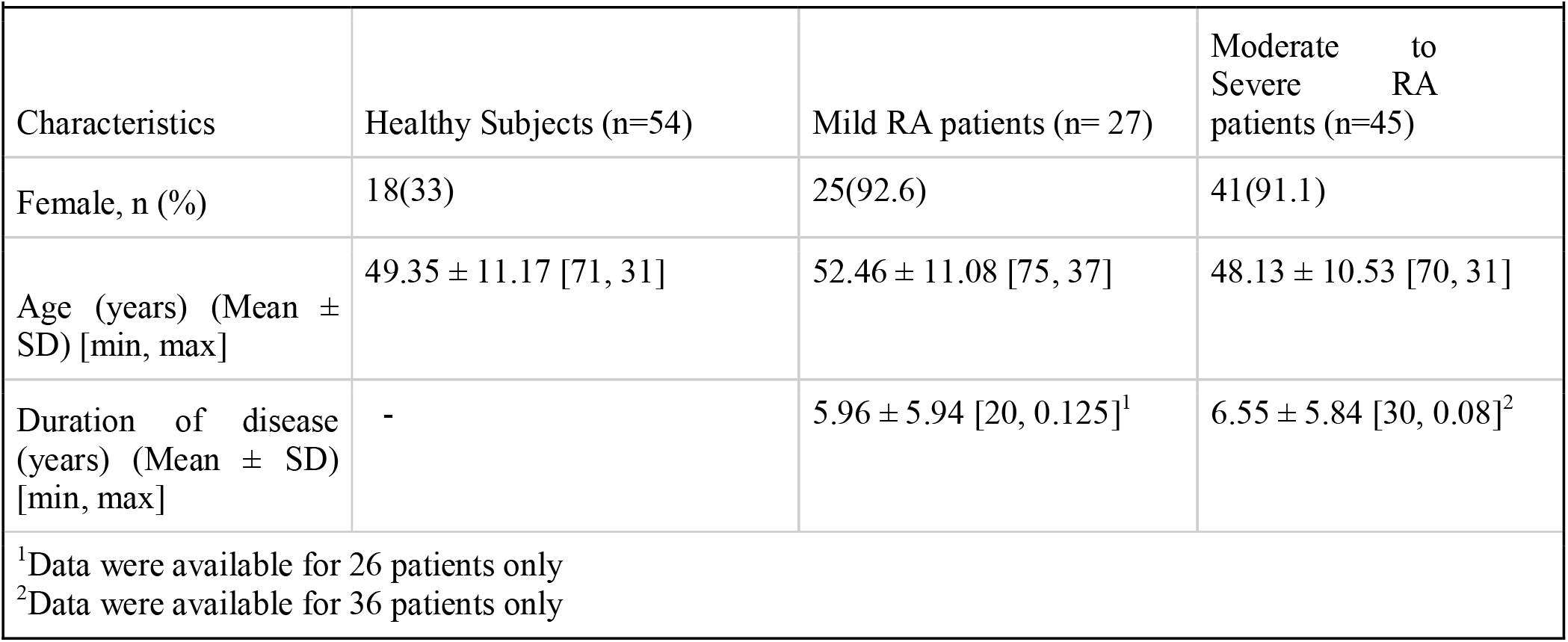
Demographic features of Healthy subjects and subgroups of RA patients.

|  |  |  |  |
| --- | --- | --- | --- |
| Characteristics | Healthy Subjects (n=54) | Mild RA patients (n= 27) | Moderate to Severe RA patients (n=45) |
| Female, n (%) | 18(33) | 25(92.6) | 41(91.1) |
| Age (years) (Mean ± SD) [min, max] | 49.35 ± 11.17 [71, 31] | 52.46 ± 11.08 [75, 37] | 48.13 ± 10.53 [70, 31] |
| Duration of disease (years) (Mean ± SD) [min, max] | - | 5.96 ± 5.94 [20, 0.125]<sup>1</sup> | 6.55 ± 5.84 [30, 0.08]<sup>2</sup> |
| <sup>1</sup>Data were available for 26 patients only |  |  |  |
| <sup>2</sup>Data were available for 36 patients only |  |  |  |

### ELISA analysis of HIP1 Antigen in RA patient sera

HIP1 sera levels were analyzed in duplicates using sandwich ELISA. HIP1 concentration in 59 RA patients and 51 healthy controls were compared, and the median values were comparable between the two groups (0.629 ng/ml vs 0.681). The low sera concentration is expected of a cytoplasmic protein. **(Fig. 1)**

**Fig.1.**
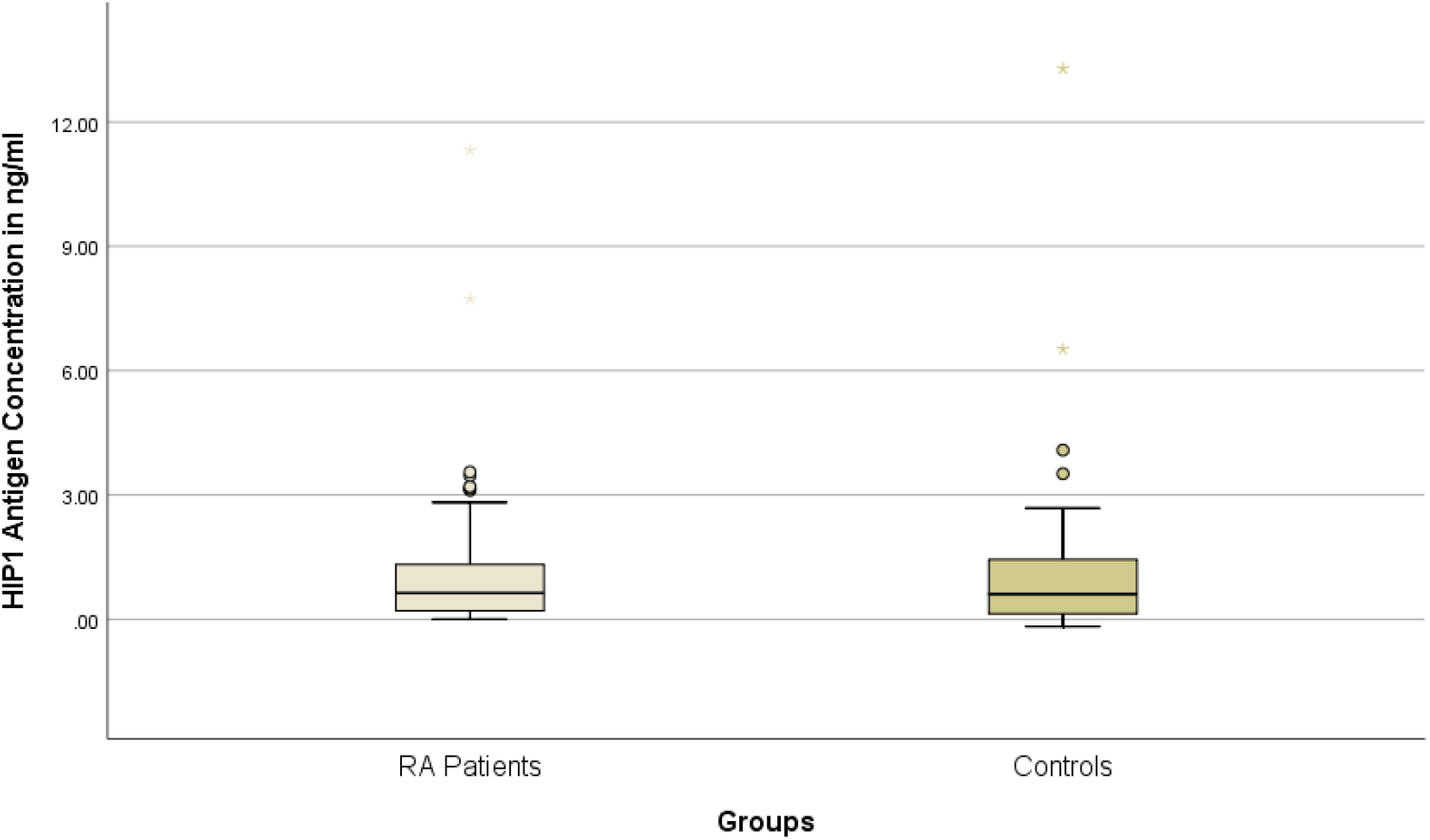
Serum HIP1 Antigen concentration in RA Patients and age matched controls. Boxplot showing Concentration of HIP1 antigen in ng/mL in Rheumatoid patients(n=59) and control samples (n=50). The X-axis represents RA patient group and control group, whereas the Y-axis represents HIP1 antigen concentration in ng/ml. The dark horizontal line on the box represents the median value (0.629 for the RA group & 0.681502 for control). The box represents the interquartile value. The difference between RA patients and control was not significant (p value=0.762, Mann Whitney Test).

### Autoantibodies to HIP1 in RA patient sera

In the absence of a commercially available kit, an immune response against HIP1 was explored using an in-house developed ELISA. Sera from 72 RA patients and 54 age-matched controls were used in duplicates to detect autoantibodies against HIP1. The median absorbance values differed significantly between the two groups, with higher HIP1 auto-antibodies in RA patients (0.32) as compared to healthy controls (0.19) (p<0.05) (**Fig. 2**).

**Fig. 2.**
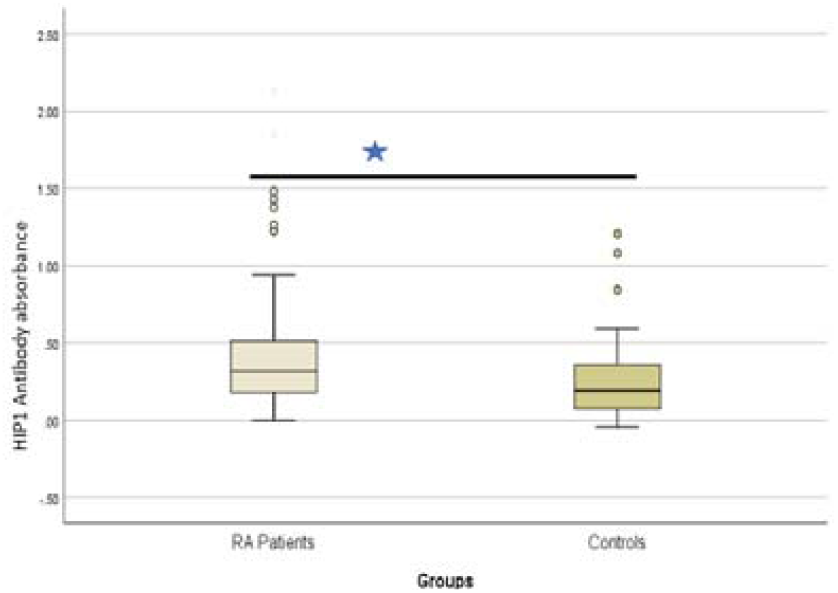
Serum HIP1 Antibody absorbance values in RA Patients and age matched controls. Boxplot showing HIP1 antibody absorbance values in RA patients (n=72) and control samples (n=54). The dark horizontal line on the box represents the median value (0.32 for RA patients and 0.19 for controls). The box represents the interquartile value. The difference between RA patients and control was significant (p value=0.002, Mann-Whitney Test).

An ROC curve for this ELISA yielded 69% sensitivity and 59% specificity, and the AUC value was 0.659 with a 95% confidence interval ranging from 0.563-0.755. The accuracy was 57.14% in the validation set when using 0.2226 as the cut-off value for the absorbance values. This assessment yielded significantly more RA patients with positive scores (69.4 % in RA patients vs 40.7% in controls) (p<0.05) (**Fig. 3)**, thus validating the potential of ELISA to distinguish RA patients from controls.

**Fig. 3.**
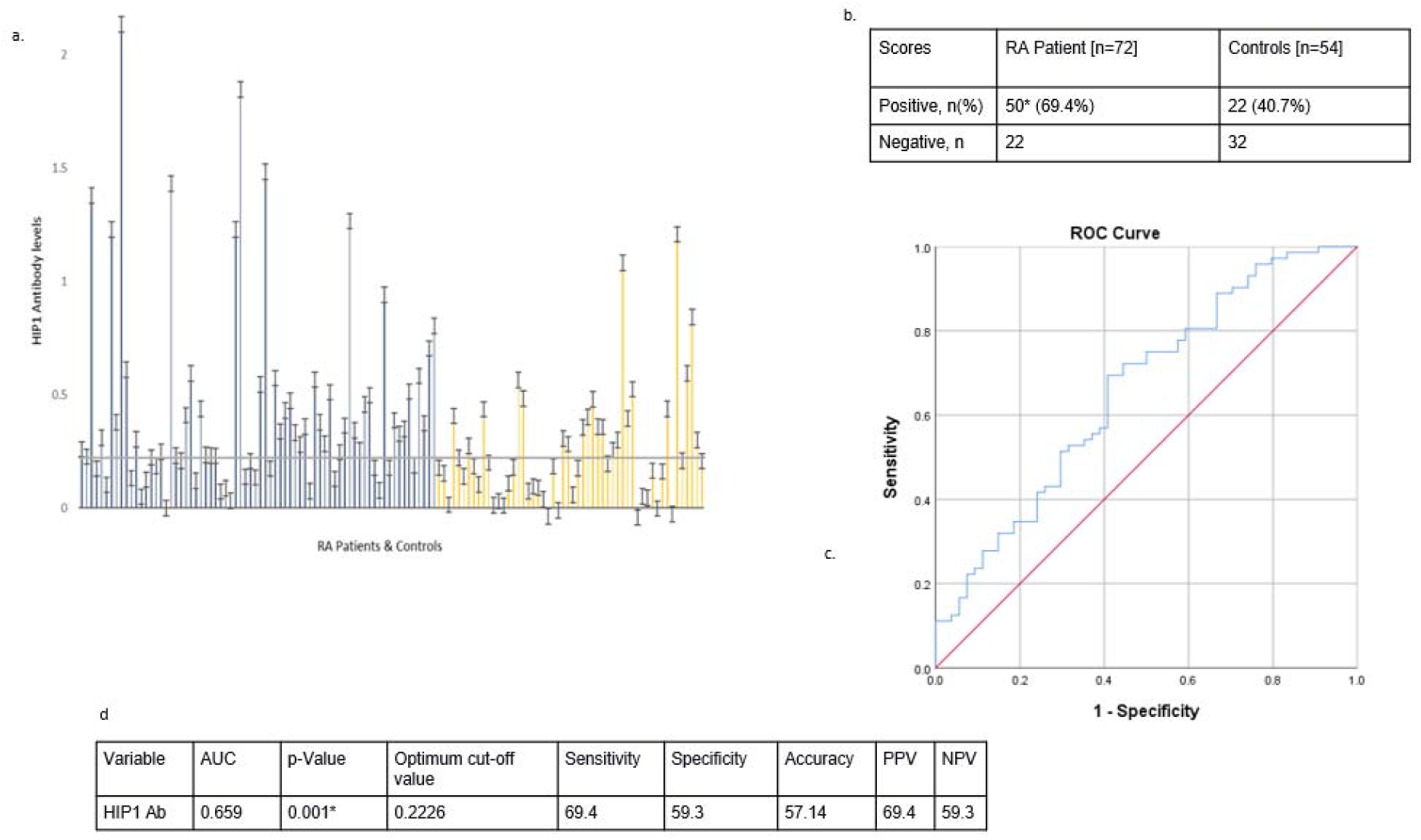
ROC analysis of HIP1 autoantibodies in RA patients and age-matched controls. (a) average absorbance (ELISA values) with their standard deviations for 72 RA patients (represented as blue bars) and 54 age-matched controls (represented as yellow bars). (b) numbers of positives in the RA patient group and age-matched control group (c) ROC curve for the predictive value of serum concentration of HIP1 in RA vs age-matched healthy controls, (d) AUC and validity of HIP1 Ab to differentiate between RA patients and the control group. (Note: Sensitivity, specificity, accuracy, PPV and NPV are in percentages. AUC: area under the curve; NPV: negative predictive value; PPV: Positive predictive value) *Difference between RA & control’s positive & negative values is significant (p=0.001 Pearson’s Chi-square test)

Further, RA patients were categorized into two subgroups: (a) mild (n=27) and (b) moderate to severe cohorts (n=45) to assess any correlation of HIP1 antibody levels with disease activity. We found significantly higher HIP1 antibodies among the moderate to severe cohort (0.35) in comparison to the mild cohort (0.25) (p<0.05) (**Fig. 4**). Similarly, ROC analysis for the same yielded 63% sensitivity and 63% specificity, and AUC was 0.644 with a 95 % confidence interval from 0.521 to 0.777 (**Fig 5)**. The accuracy was 63.38% in the external validation set using 0.2936 as the absorbance cut-off value. Thus, among the RA patients, the percentage positivity in the moderate to severe group was significantly higher (63.5%) vs those in the mild group (37%) (p<0.05).

**Fig. 4.**
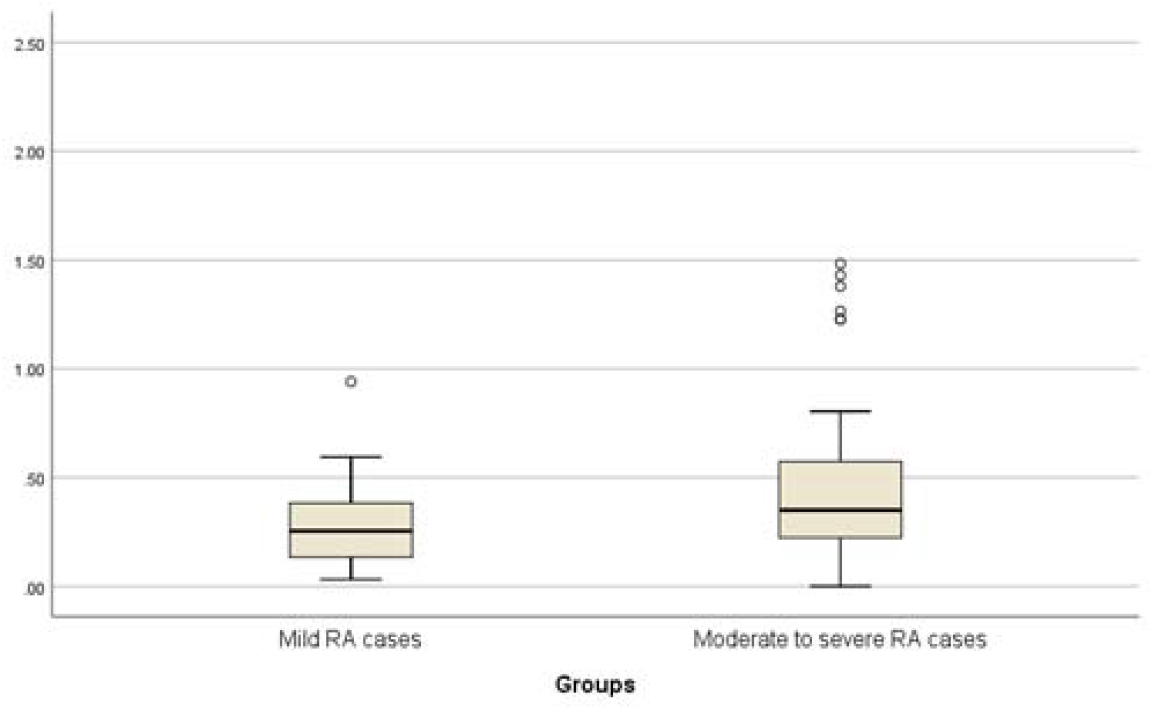
Box plot - Serum HIP1 Antibody Absorbance value in subgroups of RA Patients. Boxplot showing HIP1 antibody absorbance value in two subgroups: Mild (n=27) and Moderate to severe (n=45). The X-axis represents the two subgroups, whereas the Y-axis represents the HIP1 absorbance values. The dark horizontal line on the box represents the median value (0.25 for mild RA patients and 0.35 for moderate to severe RA patients). The box represents the interquartile value. The difference between Mild groups and Moderate to severe group was significant (p value=0.042, Mann-Whitney Test).

**Fig. 5.**
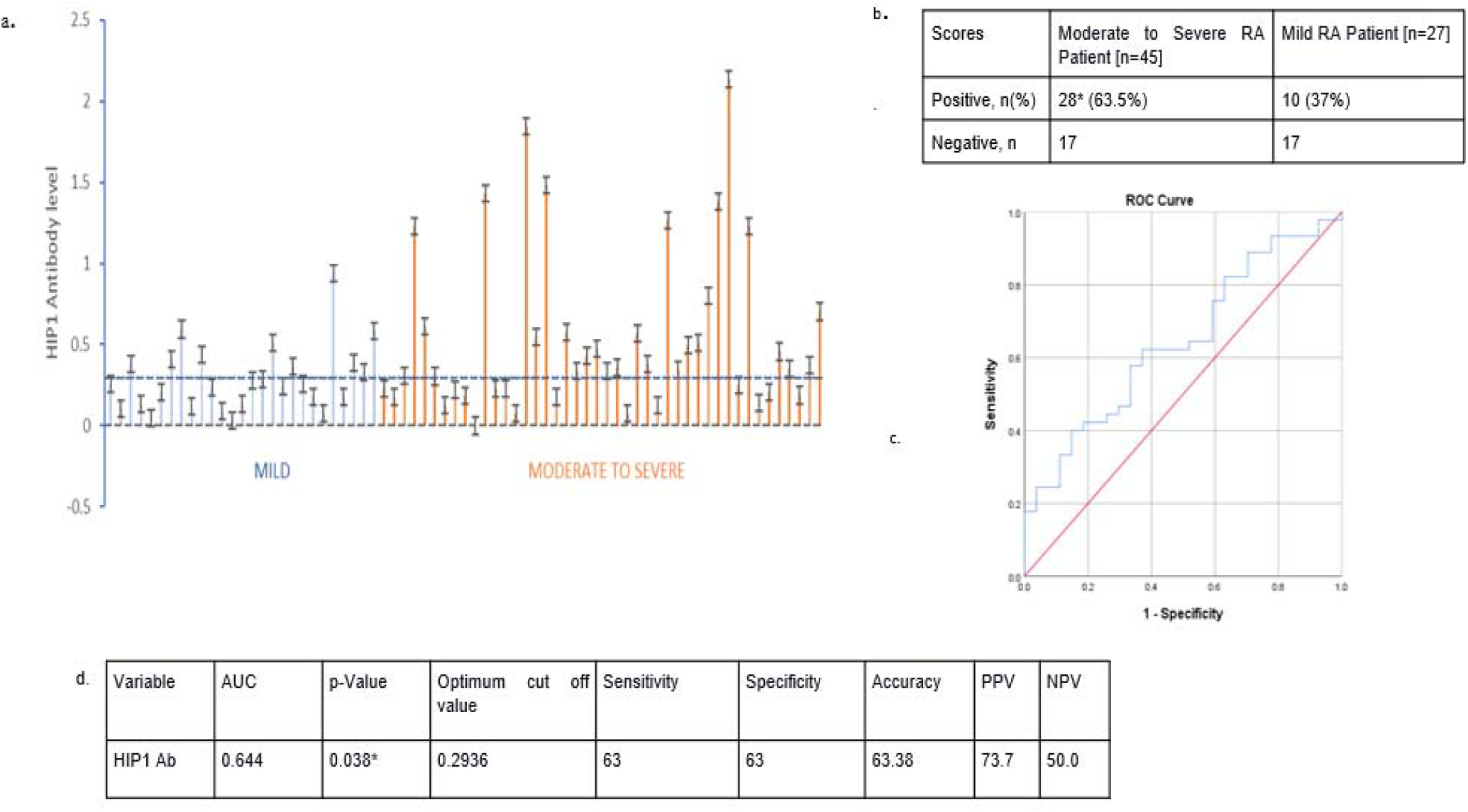
ROC analysis of HIP1 autoantibodies in subgroups of RA patients. (a) average absorbance values with their standard deviations for 27 mild RA patients (represented as blue bars) and 45 moderate-to-severe RA patients (represented as orange bars), (b) numbers of positives in subgroups of RA patients (c) ROC curve for the predictive value of serum concentration of HIP1 in RA subgroups, (d) AUC and validity of HIP1 Ab as a severity biomarker to differentiate between subgroups of RA patients. (Note: Sensitivity, specificity, accuracy, PPV and NPV are in percentages. AUC: area under the curve; NPV: negative predictive value; PPV: Positive predictive value) *Difference between subgroups of RA patient’s positive & negative values is significant (p=0.038 Pearson’s Chi-square test)

### Cytokines of Th1/Th2/Th17 lymphocytes in a subset of RA patients

The cytokine levels of Th1/Th2/Th17-related lymphocytes were explored in a subset of RA patients and Controls. Although we found no significant differences in levels of IL-2, IL-4, IL-10, IFN-gamma and TNF alpha, IL-6 levels were significantly raised in RA patients (3.96 pg/ml) as compared to controls (1.76 pg/ml) (p=0.047) and correlates with disease progression **(Fig. 6a & b)**. Further, levels of IL-6 and combined Th1(IL-2, IFN-gamma) cytokines correlated positively with disease progression **(Fig. 7a)**. Additionally, a significant positive correlation was seen between the pro-inflammatory cytokines, IL-6 and Th1 cytokines exclusively in the RA patient group (p<0.05) **(Fig. 7)**.

**Fig. 6.**
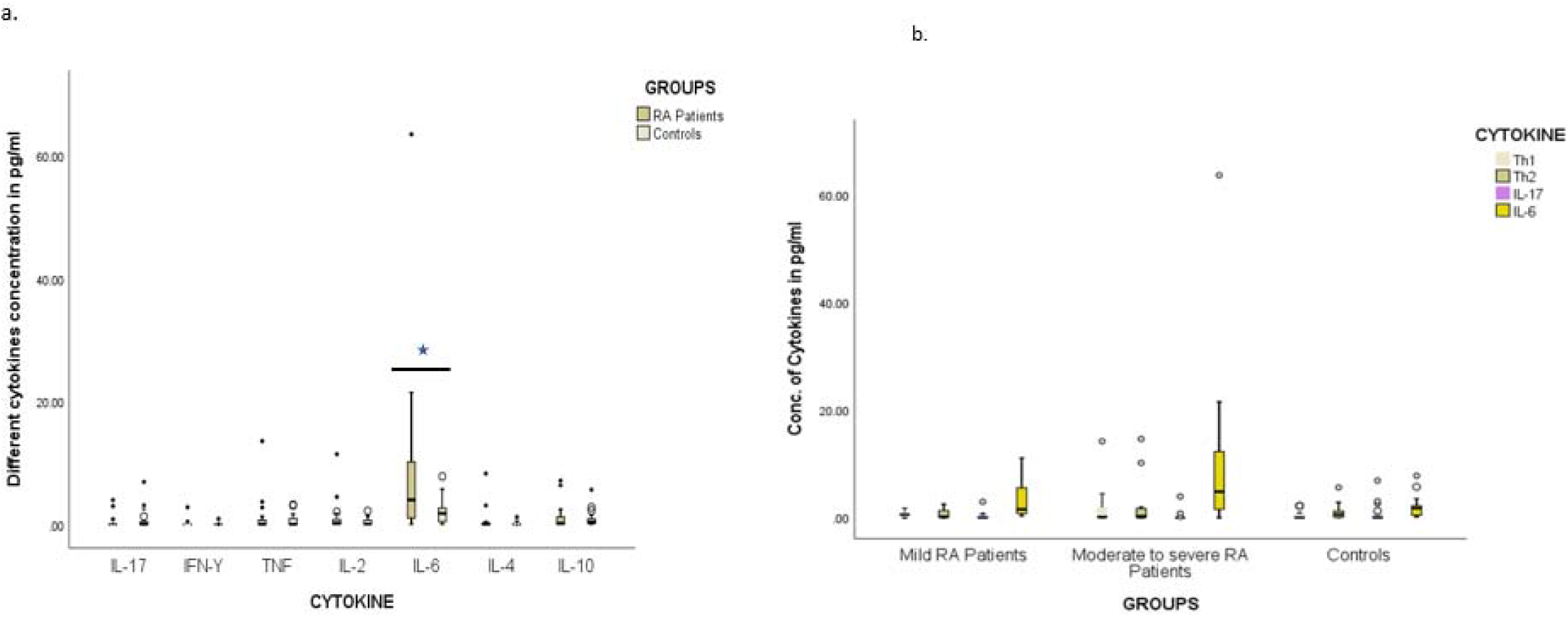
Box plot - Serum Th1/Th2/Th17 Cytokines value in RA Patients and Controls. (a) Boxplot showing concentrations of (pg./mL) IL-17, IFN-G, TNF, IL-2, IL-6, IL-4, IL-10 cytokines secreted by Th1/Th2/Th17 T cells in RA patients(n=23) and control samples (n=19). The X-axis represents different cytokine (IL-17, IFN-G, TNF, IL-19, IL-6, IL-4, IL-2) types in the Control group and in the RA group, whereas the Y-axis represents the Cytokine concentration in pg./ml. The boxes represent the interquartile values. (b) Boxplot showing concentrations of (pg./mL) Th1(IFNY, IL2), Th2(IL-10, IL-4), IL-17 and IL-6 cytokines in subgroups of RA patients and controls.

**Fig. 7.**
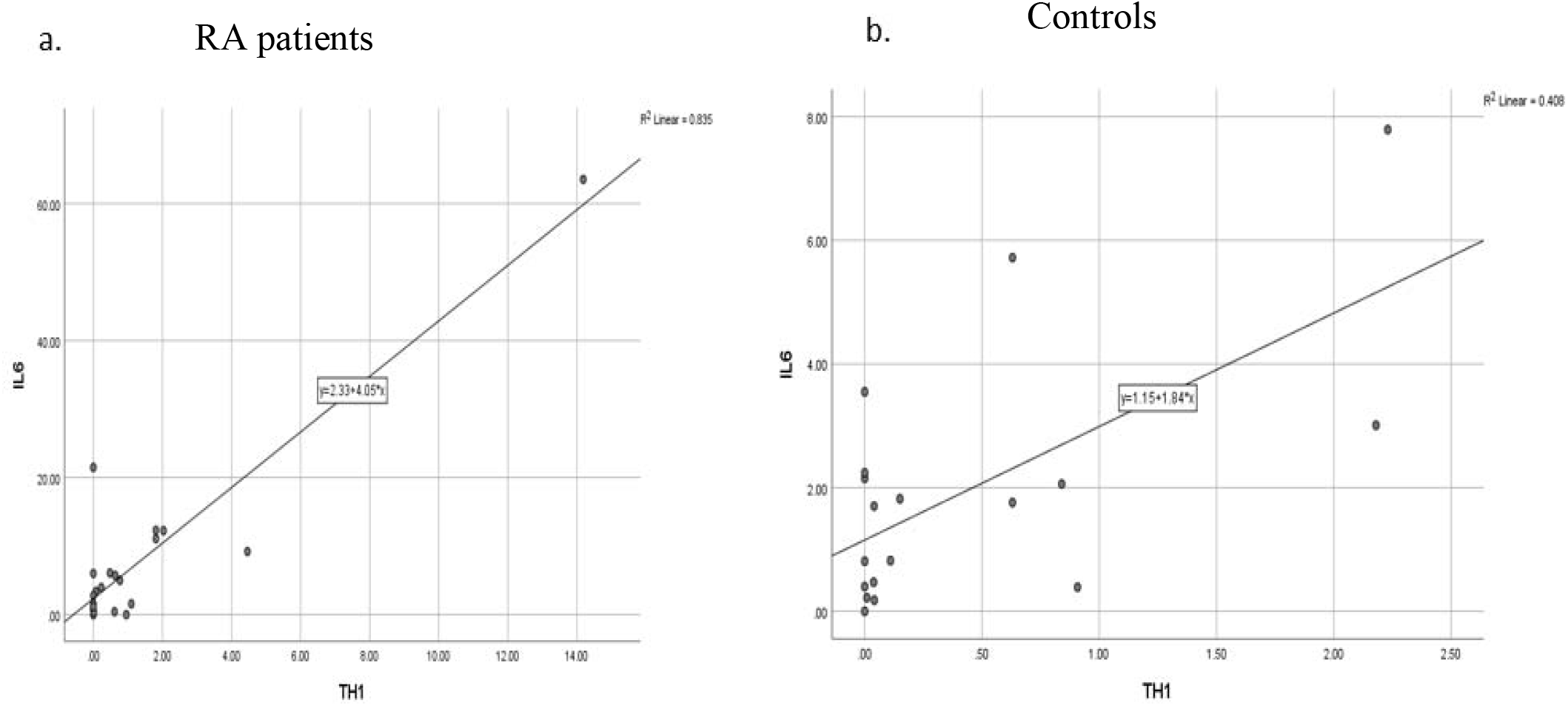
Correlation between IL-6 and Th1 levels in RA patients and healthy controls. (a) Correlation between IL-6 and Th1 levels in RA patients (p=0.024 Spearman’s Rho test) (b) Correlation between IL-6 and Th1 levels in healthy controls (not significant).

## Discussion

This is the first report demonstrating a substantial fraction of RA patients with higher autoantibody reactivity against HIP1 as compared to healthy controls (p=0.002). We detected these autoantibodies using an in-house developed ELISA, and used purified 3’HIP1 as antigen, as reported earlier [13]. The AUC for ROC of this autoantibody ELISA is 0.659, with an accuracy of 57.14%, 69% sensitivity and 59% specificity. The AUC can be further optimized by increasing the sample size. However, it is noteworthy that the technique was still able to predict significantly more numbers of positives (69.4%) in RA patients as compared to controls (40.7%). Further, HIP1 autoantibody detection had higher accuracy (63.38%) when compared between two subgroups of patients (mild vs moderate/severe disease) indicating its potential to distinguish mild disease from moderate/severe disease. However, there are few limitations, which need to be considered. One is that we are detecting autoantibodies only against the ‘C’ terminus and not the full length HIP1 protein, indicating probability of some loss of reactivity. Also, modification of many proteins by citrullination in RA is one of the reasons for a hyperactive autoimmune response [23-24]. Thus, it would be pertinent in future to explore anti-citrullinated antibody response against HIP1.

We observed similar HIP1 levels in sera of RA patients and controls. This is expected based on HIP1 being a cytosolic protein and is corroborated in other studies in cancer patients [13]. Higher HIP1 expression mediated synovial fibroblast migration is reported in rat model of RA [11,16,25-26]. This implicates measuring synovial fluid HIP1 and its autoantibodies to be a more direct and better indicator of their local expression from FLS cells, however, we considered here a non-invasive strategy as a first step.

As chronic inflammation mediates RA progression, we compared Th1/Th2/Th17 lymphocyte-related cytokine profile in sera of a subset of RA patients and healthy controls [27]. As cytokines generally act locally, most cytokine concentrations were found almost undetectable in both patients and controls. However, the proinflammatory Th1 cytokines (IL-2 and IFN-γ) and IL-6 together were significantly raised in RA patients, more so in the active disease subgroup. Also, a significant positive correlation was observed between IL-6 and Th1 cytokines exclusively in RA patients (p=0.024). IL-6, a key cytokine involved in chronic inflammation is also induced in collagen-induced arthritis in experimental animals [28]. Previous studies have also highlighted the role of IL-6 in inducing immunological abnormalities and thrombocytosis, which give rise to joint and systemic inflammation associated with RA and are found elevated with increasing disease activity in RA [29-30]. The correlation between Th1 and IL-6 specifically in RA patients is relevant owing to their proinflammatory nature and warrants further investigation as prognostic indicators of disease response in a larger cohort.

Thus, our study demonstrates that HIP1 autoantibody levels are higher in active RA patients. HIP1 may thus serve as a prognostic non-invasive biomarker. Also, combination of Th1 (IFN-gamma, IL-2) and IL-6 should be explored to assess their prognostic ability, as they show exclusive positive correlation in patients. Various studies have explored biomarkers, such as Clusterin, S100A8/A9, 14-3-3η, Galectin-1, Calprotectin, in RA disease. However, the need for a specific and sensitive marker/s is still unmet. Thus, it could be useful to create a combination of few promising markers to stratify patients with active disease, and identify the most sensitive and specific combination in future.

## Methods

### Serum Sample Collection & Storage

Sera were collected from 72 RA patients and 53 age-matched healthy controls. RA patients were recruited from the Department of Rheumatology, Sir Ganga Ram Hospital, New Delhi. Healthy controls were recruited from the Department of Preventive Health & Wellness, Sir Ganga Ram Hospital, New Delhi (January 2021 to June 2022). The studies involving human participants were reviewed and approved by the Medical-ethics committee of Sir Ganga Ram Hospital (EC/07/19/1558). All the patients/participants provided their informed consent to participate. **We confirm that all experiments were performed in accordance with relevant guidelines and regulations**.

5-10 mL of blood was collected in serum separator tubes (SST) vials. The vials were kept at room temperature for 30 minutes to one hour and centrifuged at 2000 RPM at 24□. Supernatant/Serum was carefully pipetted out and stored in microcentrifuge tubes at -80□.

#### Inclusion Criteria

RA patients aged 30 or above were chosen according to the 2010 EULAR RA classification criteria.

#### Exclusion Criteria

For age-matched healthy controls, patients aged < 30 years were excluded, and those with a history of COVID-19, heart-related issues/disturbances in ECG, bone-related issue/arthralgia/osteoarthritis, infections like HIV-AIDS, pregnancy and malignancy.

### ELISA for HIP1 Antigen in Sera

ELISA was performed with a human HIP1 ELISA kit (Bio String’s, Pennsylvania, USA) and as per the manufacturer’s instructions.

### In-house ELISA for HIP1 Autoantibodies in sera

96 well Maxi sorb plates were coated with recombinant and purified 50uL of recombinant 3’ HIP1 antigen (5 ug/ml) [Abgene]. Standardization was performed using different blocking reagents (Casein/skim milk). Plates were washed twice with TBST and blocked with 300uL, 5% skim milk in TBST overnight at 4 degrees. 100uL sera samples (1:2 diluted in blocking solution) were added in duplicates. Plates were sealed and incubated at room temperature for 1 hour. The plates were washed five times with TBST and incubated with 50uL, 1:20,000 goat anti-human IgG biotin-conjugated secondary antibody (diluted in blocking solution) for 30 minutes. Plates were washed five times with TBST and incubated with 50uL, 1:3000 avidin-biotin complex reagent for 30 minutes and washed two times. 50uL of the 1-Step Ultra TMB substrate was added to all the wells. Plates were incubated for 30 minutes, and reactions were terminated by adding 50uL 2M Sulfuric acid. Absorbance was measured at 450 nm using an ELISA reader.

### Cytokine Bead Assay for measuring Th1/Th2/Th17 cytokines

Cytokine Bead Array (CBA Kit, BD Biosciences) for measuring human Th1/Th2/Th17 related cytokines was used to measure the concentration of various cytokines (IL-2, IL-4, IL-6, IL-10, TNF, IFN-gamma, and IL-17A) in sera samples. This was performed in flow cytometer BD FACSaria III* according to the manufacturer’s protocol.

### Statistical Analysis

Statistical analyses were performed using SPSS software version 26.0 (IBM Corp., Armonk, New York). A p< 0.05 at a confidence interval of 95% was considered significant. HIP1 antigen, antibody and cytokine levels in RA patients and controls were analyzed using a non-parametric test (Mann-Whitney Test). Receiver operating characteristic (ROC) analysis was performed using Pearson’s Chi-square test. To assess the statistical significance between Th1 and IL-6, a correlation curve was drawn using Spearman’s Rho Test.

## Data Availability

All data produced in the present study are available upon reasonable request to the authors

## Acknowledgements

We acknowledge Peeyush Prasad, Sister Manisha, and Sakshi for their help in patient sample identification and collection. We thank Parul Chugh for assistance in statistical analysis. We thank Delhi Rheumatology Association of India & SGRH for financial support. We thank Council of Scientific and Industrial Research (CSIR) for providing Junior Research Fellowship (JRF) to Ms. Surbhi.

## Author Contributions

S and AG performed experiments, analyzed the data & prepared the manuscript. SV collected samples. VC & SR provided blood samples from patients and healthy controls. VC analyzed patient’s data and helped in manuscript preparation. NKG helped in manuscript preparation. All authors read and approved the final manuscript. SM conceptualized, supervised and helped in manuscript preparation.

## Data Availability

The data generated/or analyzed during the current study are available from the corresponding author on reasonable request.

## Competing Interest

The author declares no competing interest.

## Notes

### Competing Interest Statement

The authors have declared no competing interest.

### Funding Statement

This study was funded by Delhi Rheumatology Association & Sir Ganga Ram Hospital. We would also like to thank Council of Scientific and Industrial Research (CSIR) for providing Junior Research Fellowship to Ms Surbhi.

### Author Declarations

Ethics committee of Sir Ganga Ram Hospital (EC/07/19/1558)

### Summary of Updates

Acknowledgment section is updated.

